# Corona Epidemic in Indian context: Predictive Mathematical Modelling

**DOI:** 10.1101/2020.04.03.20047175

**Authors:** Jyoti Bhola, Vandana Revathi Venkateswaran, Monika Koul

## Abstract

The novel Coronavirus pathogen Covid-19 is a cause of concern across the world as the human-to-human infection caused by it is spreading at a fast pace. The virus that first manifested in Wuhan, China has travelled across continents. The increase in number of deaths in Italy, Iran, USA, and other countries has alarmed both the developed and developing countries. Scientists are working hard to develop a vaccine against the virus, but until now no breakthrough has been achieved. India, the second most populated country in the world, is working hard in all dimensions to stop the spread of community infection. Health care facilities are being updated; medical and paramedical staffs are getting trained, and many agencies are raising awareness on the issues related to this virus and its transmission. The administration is leaving no stone unturned to prepare the country to mitigate the adverse effects. However, as the number of infected patients, and those getting cured is changing differently in different states everyday it is difficult to predict the spread of the virus and its fate in Indian context. Different states have adopted measures to stop the community spread. Considering the vast size of the country, the population size and other socio-economic conditions of the states, a single uniform policy may not work to contain the disease. In this paper, we discuss a predictive mathematical model that can give us some idea of the fate of the virus, an indicative data and future projections to understand the further course this pandemic can take. The data can be used by the health care agencies, the Government Organizations and the Planning Commission to make suitable arrangements to fight the pandemic. Though the model is preliminary, it can be used at regional level to manage the health care system in the present scenario. The recommendations can be made, and advisories prepared based on the predictive results that can be implemented at regional levels.

## Introduction

Viruses have been considered as inconsequential pathogens in humans for long in comparison to plants. The mortality caused by viruses in humans has been very low in comparison to other diseases such as Cancer, Cardio-vascular diseases and Tuberculosis (CGHR Report, 2017). However, viruses have been reported to exacerbate the symptoms and have more serious implications on human health if a person is suffering from some auto-immune disorder, infectious disease and has compromised immune system.

In India, for diseases such as Tuberculosis, HIV and Cancer, the related mortality rate is so high that researchers and funding agencies have under-estimated the implications viral diseases can have on public health and socio-economic security. Funding agencies and data scientists have overlooked flu and other human viral pathogens; and in the past years, not much research is funded to work on drug designing or medical research in the field. It is also clear from the recent trends that viral diseases are going to spread at fast pace and many novel viruses will be unearthed in near future. Climate change and deforestation is also causing surge in outbreak of viral epidemics. The changing climate is also responsible for increase in number of vectors that accelerate the spread of pathogens (Khan et al., 2019). In the last few years, WHO (World Health Organization) has been continuously emphasizing that the fast-developing nations such as India should triple their expenditure on health care to meet the SDG goals. The outbreak of the SARS-CoV-2 has raised the concerns for the Government of India, public policy makers and administration as the pandemic has implication on almost all sectors and strata of the society. It is therefore important to understand how the virus will fare in India and how efficiently the country can handle it without causing severe damage to human population.

### COVID Pandemic

The recent outbreak of the global pandemic Covid-19 has changed the perspective of everyone in the country regarding the viral disease outbreaks as it is affecting and infecting humans around in an exponential manner. This viral disease as well as the causative agent is novel entry to the viral world and hence is posing unforeseen challenges. SARS-Cov-2, commonly known as Novel Coronavirus, is a single, positive-stranded, RNA virus belonging to order Nidovirales (Cascella, 2020), responsible for the Current Global Pandemic Covid-19 (Huang et al, 2020). Corona viruses show enormous diversity and are evolving fast. Some of the other viruses related to this family of Coronavirus are SARS-CoV, responsible for SARS (Severe Acute Respiratory Syndrome) (Ksiazek et al., 2003) and MERS-CoV (Middle East Respiratory Syndrome) (Peeri, et al., 2020). Both these viruses emerged from animal reservoirs to cause global epidemics with alarming morbidity and mortality (Chavang, 2007). The reservoirs of the Novel Coronavirus are mostly animals found in the wild. Scientists also believe that peri-domestic mammals may also serve as intermediate hosts (Malik et al, 2020; Mudrich et al., 2020). Besides, many spontaneous mutations that keep on happening in nature are facilitating expansion of genetic diversity (He et al., 2020). There is strong evidence that human-animal contact at live game markets plays a role and thus there is zoonotic transmission of the virus (Li et al., 2020). For long, human viruses have not been considered severe pathogens as infected people develop flu like symptoms and then get cured on their own as innate immune system triggers antibody formation that provides resistance against the diseases (Chiu, 2013; Kistler et al., 2007; Wrammert et al., 2008). In both developed and developing countries the signs of common flu have not been a cause of concern though some vaccines have been developed recently and elderly are advised to take shots yearly as they have compromised immune systems (Voordouw et al., 2004). However, the spread of Novel Covid-19 has alarmed people all over the world. It is important to understand how the virus will fare in the alien environments. Therefore, interdisciplinary research involving biologists, data scientists, mathematicians, clinicians is required in order to work towards stopping the spread of these diseases and design appropriate methods, drugs to contain it before the situation gets out of hand.

The present virus that first manifested in Wuhan is different from other Corona viruses known to humankind, and that is the reason for being cautious. Basic symptoms resemble the normal flu-like symptoms that can result in cough, cold, headache and body ache. The more severe consequences include acute respiratory tract infections that culminate into pneumonia (WHO report, 2020). In more severe cases, especially old people also develop secondary infections that start affecting other vital organs and in worst cases leading to death of any individuals which can be cured for most infected individuals (Xu et al., 2020). The fast pace at which it is spreading from human to human contact is the current major reason of worry, and this what we show through a concise mathematical model in this paper. The infections can spread through air (if the infected individual is less than one meter apart from uninfected individuals), mostly through the droplets of infected people since the virus stays alive in droplets on the surfaces for many days. As soon as it gets into the host, it replicates in the body, and the body becomes a reservoir (Rothan and Byrareddy, 2020). However, in some cases, the virus can stay latent inside the body and may not cause any disease symptoms, but if this person is a carrier, then he/she can spread the disease to others who come in contact with the droplets of the person that are released during coughing or sneezing. The symptoms resemble people who catch seasonal flu like symptoms ranging from cough, cold, fever to shortness of breath (WHO, 2020; Wu, 2020). As the symptoms shown by the person infected with Novel Covid-19 are similar and over lapping with other common flu, it is not easy to identify the carriers and therefore, the transmission cannot be easily contained.

First few reported cases of Covid-19 infected people had shared a history of human-animal contact at live game markets. The World Health Organization has declared it a pandemic after more than **200**,**500** confirmed cases of infection and more than 9,000 deaths across 115 countries (https://www.who.int/).The virus epidemiology has been understood in China where research teams, institutes and scientists and various cross sectors worked in coordination and today China has been able to control it in the worst affected areas (Zhau et al., 2020; Zhang et al., 2020). This shows the importance of coordinated, well-funded research, especially in fields crucial to humankind’s survival.

India tops the list of countries with highest population and has been trying hard to negotiate the problems and challenges thrown by this overwhelming population (Ministry of Health, GOI, 2020). The rise in population has already created crisis for health sector, education and has caused poverty elevation. With the outbreak of the novel Coronavirus Covid-19, the country must gear up to confront it. It is important for Indian Scientists to come together and study the pathogenicity in the Indian context. In India, the first case was reported in the end of January this year, and the number has grown to touch 800 as on March 28, 2020. But India is a peninsula and the temperature, humidity and topography is variable, hence the factoring of variables will also have an implication on mortality and morbidity. Also, education, awareness and understanding of people, socio-economic status is variable, so the infection percentage, the magnitude of impact will also be different. This calls for regional data assessment and modelling. Besides, mathematical and ecological modelling can help in predicting the disease course. The data generator through these models can consider various variables that are specific to the country and give some predictions. This can help in a fair assessment and put all the fake and unscientific assumptions on hold. This data can also help in giving recommendations to the health care agencies.

The present article aims to throw light on the present situation in India and support the action plan of physical distancing as stressed upon by GoI through a mathematical model capturing the spread of the disease. The emergence of yet another outbreak of human disease caused by a pathogen from a viral family formerly thought to be relatively benign underscores the perpetual challenge of emerging infectious diseases and the importance of sustained preparedness. Major gaps in our knowledge of the origin, epidemiology, duration of human transmission, virus evolution, and clinical spectrum of disease need fulfillment by future studies. Research teams including biologists, biotechnologists and medical practitioners all over the world are trying to understand biology, etiology and epidemiology of the diseases (Novel, C. P. E. R. E., 2020; Lipsitch et al, 2020; Rothan and Byrareddy, 2020). Mathematicians, data scientists and statisticians have proposed various models to decipher and predict the behavior of the epidemic in various environments (Mandal et al, 2020; Nesteruk, 2020; Ming et al, 2020; Maier and Brockmann, 2020). Scientists have also been using dynamic bipartite graphs to model the physical contact patterns that result from movements of individuals between specific locations based on the trends available each day (Zhao, 2020)

We used the software Mathematica to do predictive modelling based on the data that is presently available on the mortality due to the Pandemic Spread by Covid-19.

In the present context, population of India can be classified into three broad classes; namely

*I(t)*: the number of infectives

*S(t)*: the number of susceptible

*R(t)*: the number recovered (recovered, died, or naturally immune to the disease)

Clearly, *I(t)+S(t)+R(t) = N* where *\*N* is the total population of India (\**N* is itself a variable, but treated as a constant in this model consistent with the fact that the course of this epidemic is short compared with the lifetime of an individual).

This can be seen as a three-compartment model (Figures 1 and 2) and what is our interest is to have the minima of the *I(t)* compartment under the prevailing circumstances.

**Figure 1.**
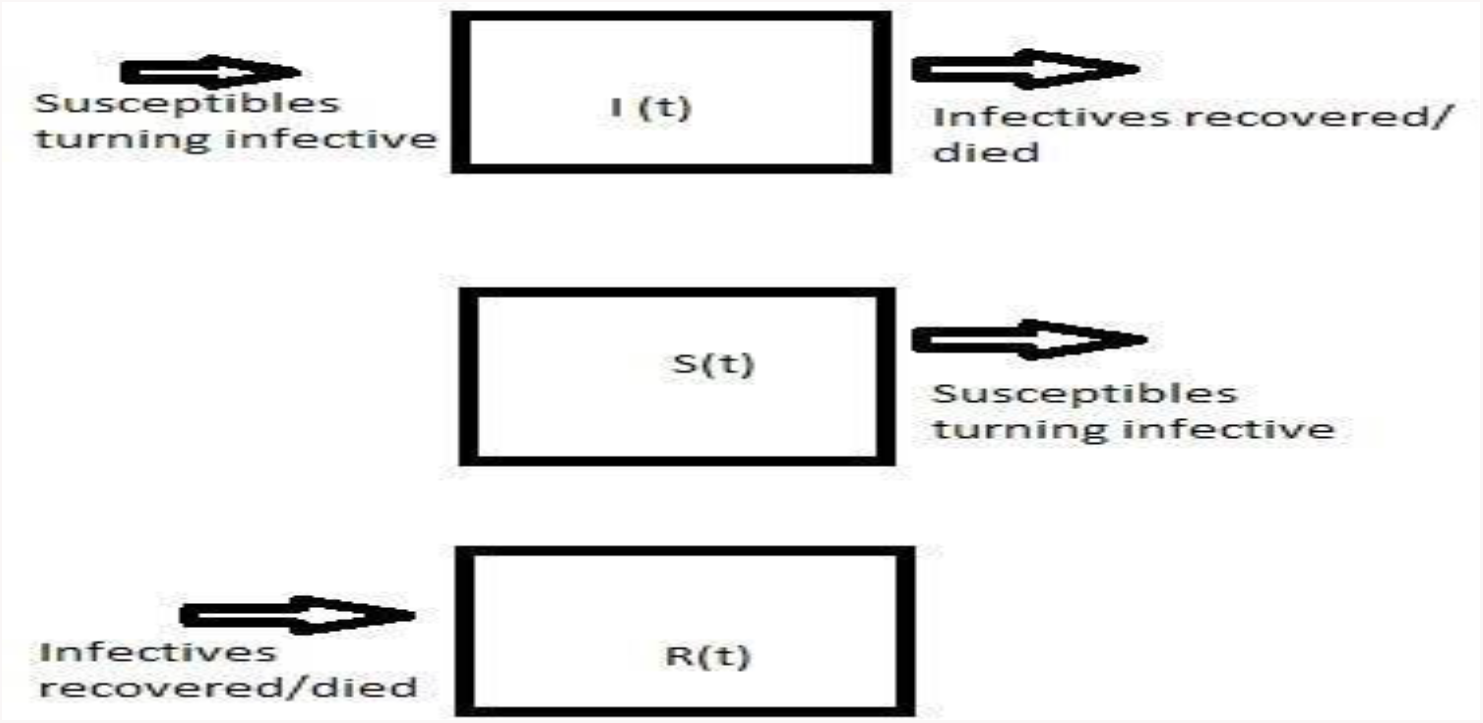

**Figure 2.**
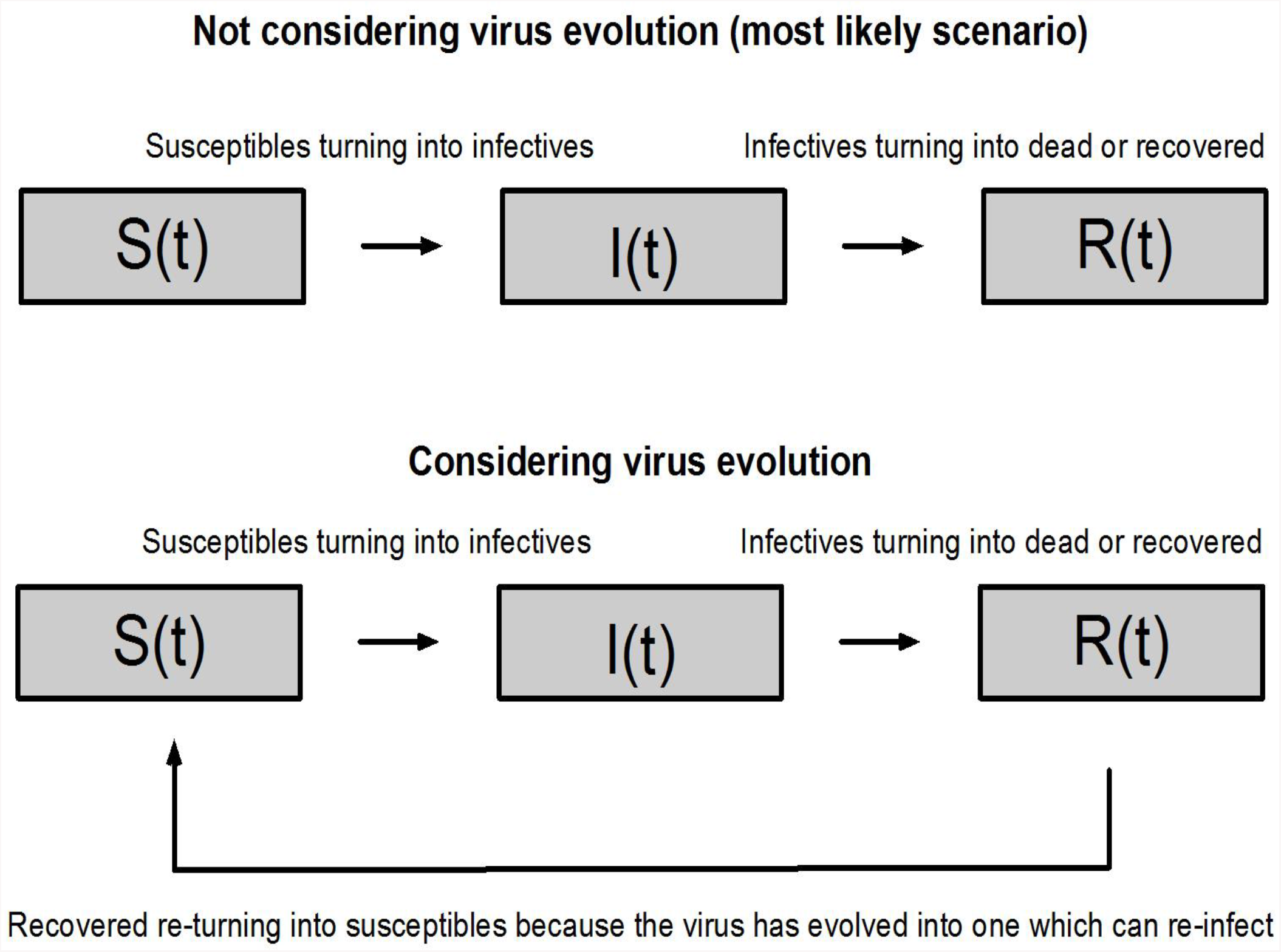

From what is known about corona viruses, it is evident that the per capita rate of increase in the number of infectives is directly proportional to the number of susceptible in the vicinity of an infective and hence, the total intake in the first compartment looks like *(kS)I* ; where *k* signifies the rate of transmission indicated by the average number of people who will catch the virus from one infected person.

The basic differential equation system [SIR-Model] (Barnes and Fulford, 2002) that captures the problem is given by:

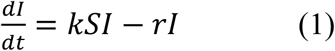

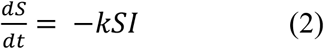

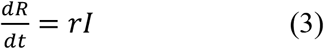

where *r* is the rate at which infectives recover or die; and clearly, these individuals can no longer remain infective. Looking at the statistics worldwide (https://www.who.int/) the value of *k* is somewhere between 1.4 and 2.5. For the sake of visualization let us take the total world population to be 100 and a single infective to begin with. A plot code in Mathematica for *k=2* (which is much lower than the actual *k* for many countries at present) gives a striking sketch (Figure 3).

**Figure 3.**
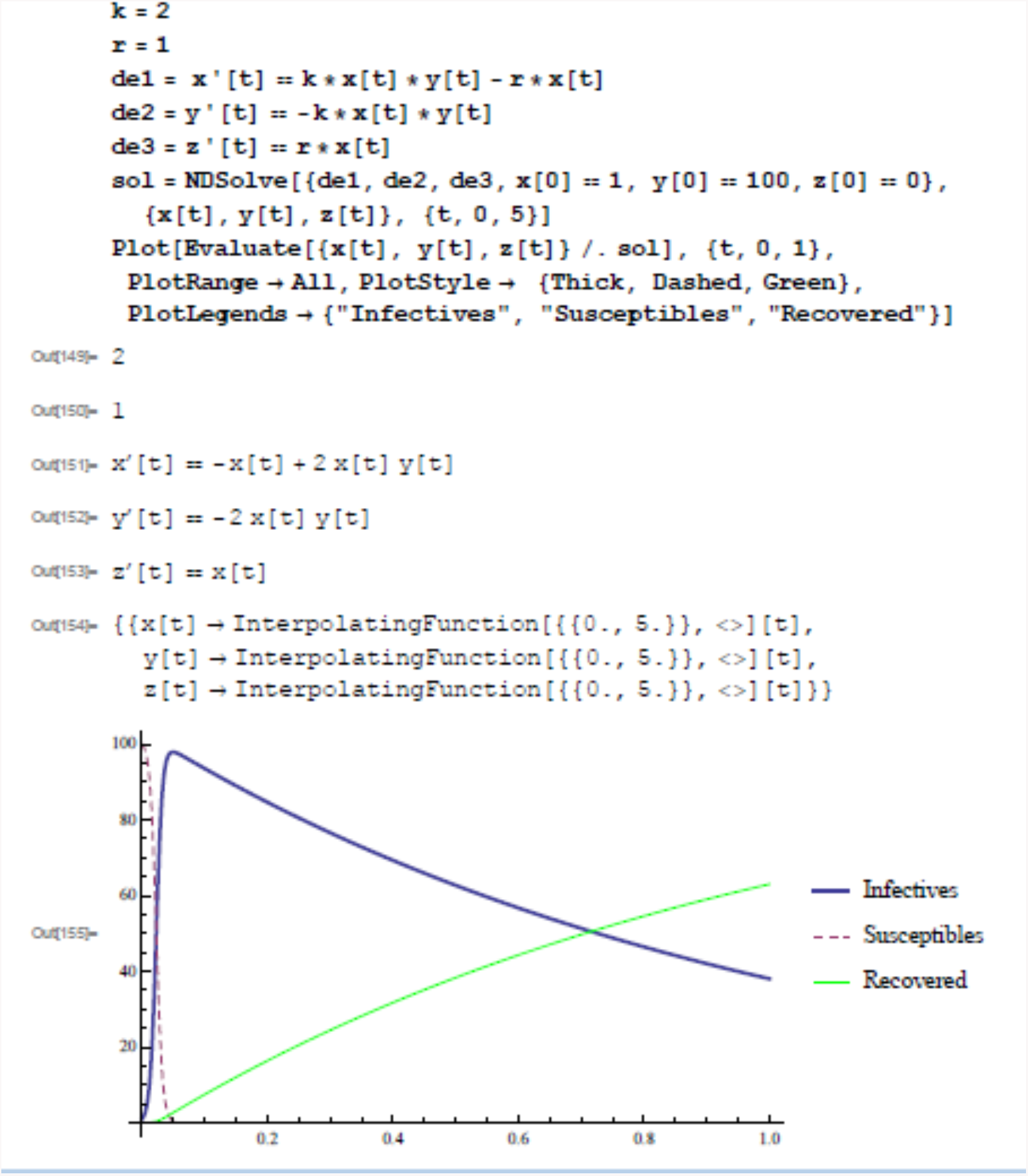

Starting with just a single infective, the infection peaks up to almost the total population size before starting to fall down. The situation in India is summarized in the https://www.mohfw.gov.in/ website.

At present, community transmission has not been validated and the *k* value is significantly less than 1 in India. The infection will still continue growing initially before attaining a peak lower than the total susceptible population this time, as indicated by Figure 4.

**Figure 4.**
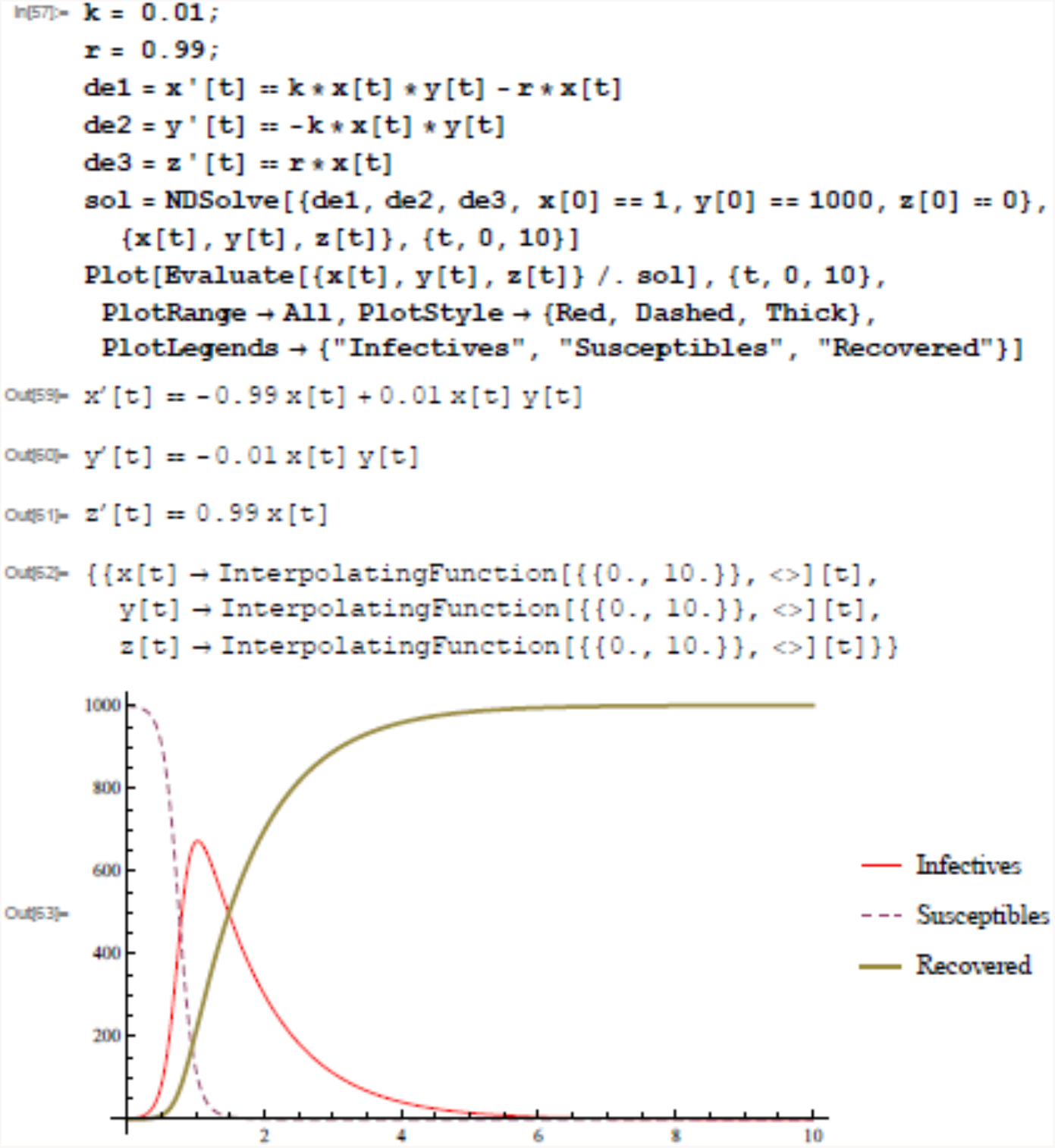

To contain the transmission of this virus, it is extremely important to contain the value of *k*. This *k* value is supposedly dependent on many factors that involve both natural (temperature, humidity) and non-natural factors or personal provisioning measures such as physical distancing, infectives wearing masks, good hygiene practices such as washing hands with soap for 20 seconds, and so on.

If *k* is greater than 1, then the disease will grow exponentially after a critical stage and become an epidemic. The exponential growth occurs because every infected individual replaces himself or herself by more than one new infected person on an average.

Scientists classify this value as R_0_ or basic reproduction number. The R_0_ for measles is around 12, the R_0_ for COVID19 is around 2.6, and for seasonal flu it is around 1.3. Figure 5 shows how the number of new cases (per transmission) for seasonal flu is negligible as compared to COVID19.

**Figure 5.**
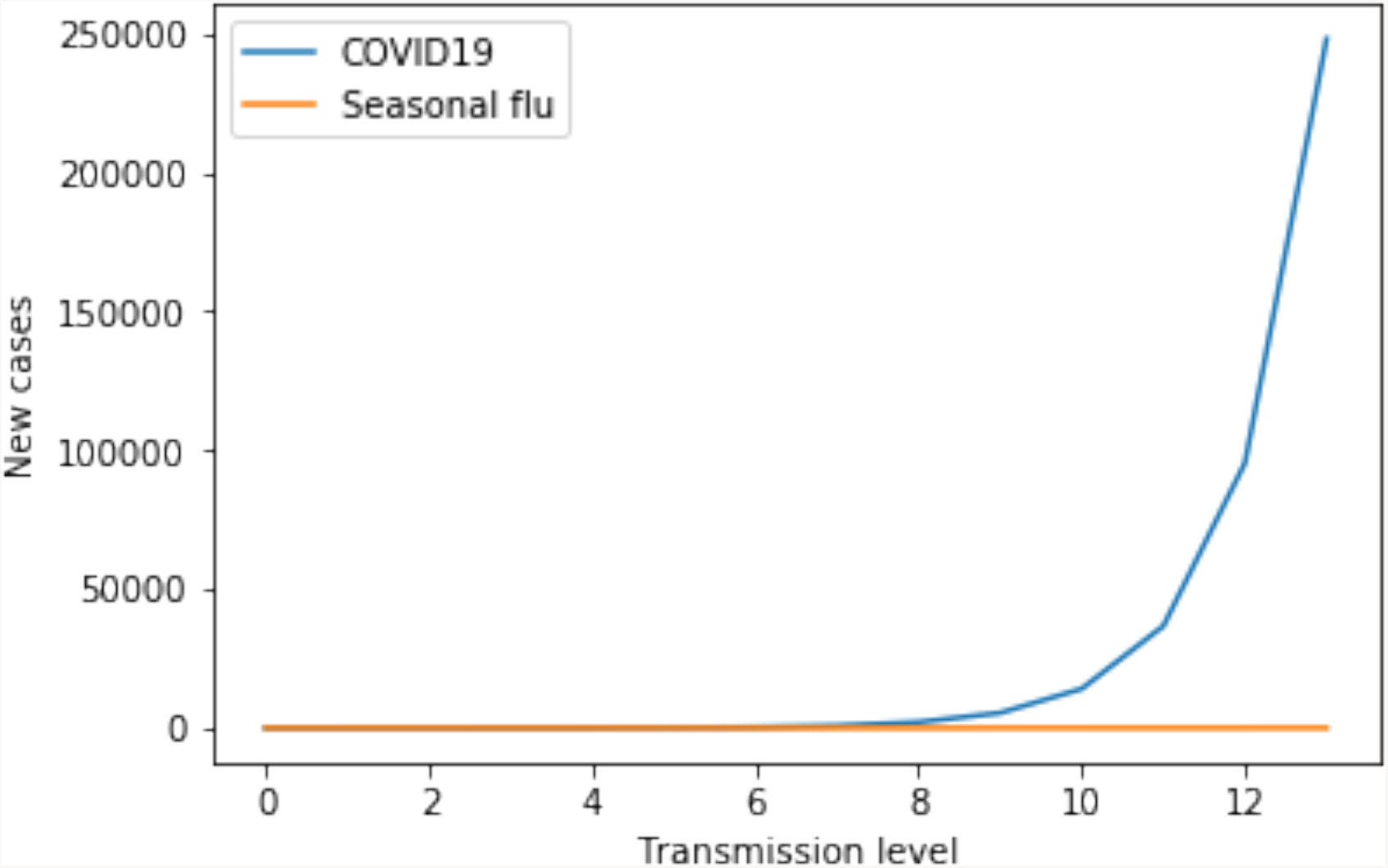

However, the exponential growth process can only continue 1) if there are sufficiently many susceptible individuals available. Once a larger fraction of the population has gone through the infection and has become immune, the probability of an infected person transmitting the infection decreases. But as shown in the previous figures, in every transmission from person to person, the numbers increase exponentially, and we would not have enough hospitals beds, medical staff and equipment to treat all infected individuals if exponentially increasing number of people have to be admitted in hospitals every day! We cannot risk such a situation because any healthcare system (of even the richest and most developed country) will breakdown and we would have unnecessary deaths. As humans, we have the knowledge to overcome such situations. We have the knowledge of science, and we should use this to avoid unnecessary deaths. This is where the second option becomes important to practice, that is 2) physical distancing to ensure we do not spread (or receive) the virus to (or from) other people. This way, the virus surviving within the bodies of already infected individuals can no longer survive by jumping to other persons. The viruses would stop surviving in the infected persons’ bodies after its 14-day incubation period in that host. This way, we can reduce the number of deaths, and also reduce the number of infected by blocking the virus from spreading.

All this standard epidemiology is going to work in the Covid-19 case only if this new strain of virus behaves in a decent way, i.e. within our current grasp of understanding. What if the virus mutation occurs causing re-infection (Figure 2) or what if there comes a second wave of infection affecting large numbers? All these apprehensions need a more detailed analysis. Scientists are working on getting a full picture of the evolution of the virus, but it might take some time. We need to give scientists this time; we all need to give medical and healthcare workers the time to deal with new patients every day. We can deal with both these situations only by lockdown, physical distancing and personal hygiene methods. The cooperation of every citizen is required.

The sate wise scenario in India as on March 28, 2020 is tabulated as under: (Source: https://www.mohfw.gov.in/)

| **Name of State / UT** | **Total Confirmed cases** | **Cured/ Discharged /Migrated** | **Death(s)** |
| --- | --- | --- | --- |
| **Andaman and Nicobar Islands** | **2** | **0** | **0** |
| **Andhra Pradesh** | **14** | **1** | **0** |
| **Bihar** | **9** | **0** | **1** |
| **Chandigarh** | **7** | **0** | **0** |
| **Chhattisgarh** | **6** | **0** | **0** |
| **Delhi** | **39** | **6** | **1** |
| **Goa** | **3** | **0** | **0** |
| **Gujarat** | **45** | **0** | **3** |
| **Haryana** | **33** | **11** | **0** |
| **Himachal Pradesh** | **3** | **0** | **1** |
| **Jammu and Kashmir** | **18** | **1** | **1** |
| **Karnataka** | **55** | **3** | **2** |
| **Kerala** | **173** | **11** | **0** |
| **Ladakh** | **13** | **3** | **0** |
| **Madhya Pradesh** | **30** | **0** | **2** |
| **Maharashtra** | **180** | **25** | **5** |
| **Manipur** | **1** | **0** | **0** |

| Name of State / UT | Total Confirmed cases | Cured/ Discharged /Migrated | Death(s) |
| --- | --- | --- | --- |
| Mizoram | 1 | 0 | 0 |
| Odisha | 3 | 0 | 0 |
| Puducherry | 1 | 0 | 0 |
| Punjab | 38 | 1 | 1 |
| Rajasthan | 48 | 3 | 0 |
| Tamil Nadu | 38 | 2 | 1 |
| Telengana | 48 | 1 | 0 |
| Uttarakhand | 5 | 0 | 0 |
| Uttar Pradesh | 45 | 11 | 0 |
| West Bengal | 15 | 0 | 1 |

The above table shows the maximum number of Covid-19 cases in Maharashtra. Maharashtra is densely populated with Mumbai being an overcrowded city that has a population size of more than 20 million people to which this highest number is being attributed to. This has happened in Maharashtra even when the state has not been the first one to get the infection in India. The same can be paralleled with the Influenza epidemic a century ago which took a toll of estimated 10-20 million people in India alone and warns us of what lies ahead of us, if the present situation goes uncontrolled, unattended or casually attended.

**Also, from the above plots (Figures 3 and 4), though the transmission rate is very low in India now, the number of infectives is bound to increase with time. The only way to get the graph of infectives as a decreasing function of time as per the proposed model is that the interaction term *kSI* in the equation (1) tends to zero. That can only happen when infectives are totally isolated from the susceptible population. The variables will keep on changing with each passing day as the virus has just reached stage 2 of its disease cycle where there are no mass causalities and things are under control**.

## Conclusions and Recommendations

Data suggests that the country is going to see a surge in the cases. However, we have to understand that variables such as hygiene, physical distancing, staying indoors, and boosting immune system can flatten the curve. Raising awareness through various platforms including the social networking websites like Facebook and Twitter is one way of containing the disease. Each household must be reached and that can be done through vernacular language since large section of the population is still not conversant in English and Hindi. Government has already taken measures such as setting testing centres and has designated isolation blocks in hospitals. This would ease the burden on the existing ones. New makeshift health centers can also be created as they did in China, which helped them win over the battle despite the high cases there.

Also, the country-lockdown announced by the government is surely going to act as infection controller and hopefully help India attend to this new challenge in the desired form. Strict measures have been announced for people who are not following Government Advisory. It is important to boost the morale of front-line workers (such as medical practitioners and para-medical staff, nursing staff, cleaning staff and house-keeping departments of the hospitals and health care centres) who are interacting directly with the patients. People who are showing anxiety symptoms due to restrictions and lock downs need constant counselling. Psychologists, self-help groups need to offer counselling through electronic media. Indians also need to fight the menace of superstitions and myths that are being propagated regarding the cure of Corona related illness. Scientists need to gear up for the task and come forward to do collaborative research work to understand the spread, containment and eventualities of the pandemic outbreak. For this, interdisciplinary teams must work together to come out with some concrete strategy. Research teams have to develop vaccines for which funding, infrastructure, and adequate facilities are required. So, Covid-19 is a reminder that science cannot take a back seat and health care, education and research should always hold a top priority. Faith in science and scientists, and optimism is important at this juncture so that India comes out as a winner in the battle.

## Data Availability

We did not generate new data. We used data from the following websites:
https://www.icmr.nic.in/
https://www.mohfw.gov.in/
https://www.who.int/
The Mathematica code mentioned (and shown in the figures) in the main article, can be obtained from the first author on request.

## References

1. Belinda Barnes and Glenn R. Fulford, Mathematical Modelling with case studies, A differential equation approach using Maple, Taylor and Francis, London and NewYork, 2002.

2. Cascella, M., Rajnik, M., Cuomo, A., Dulebohn, S. C., & Di Napoli, R. (2020). Features, Evaluation and Treatment Coronavirus (COVID-19). In StatPearls [Internet]. StatPearls Publishing.

3. Cavanagh, D. (2007). Coronavirus avian infectious bronchitis virus. Veterinary research, 38(2), 281–297.

4. Chiu, C. Y. (2013). Viral pathogen discovery. Current opinion in microbiology, 16(4), 468–478.

5. Eubank, S., Guclu, H., Kumar, V. A., Marathe, M. V., Srinivasan, A., Toroczkai, Z., & Wang, N. (2004). Modelling disease outbreaks in realistic urban social networks. Nature, 429(6988), 180–184.

6. He, X.; Yue, W.; Yan, J. SNV Reoccurrence in Multiple Regions in the Genome of 2019-nCoV. Preprints 2020, 2020020132 (doi: 10.20944/preprints202002.0132.v2).

7. https://www.google.co.in/

8. https://www.icmr.nic.in/

9. https://www.mohfw.gov.in/

10. https://www.who.int/

11. Huang, C., Wang, Y., Li, X., Ren, L., Zhao, J., Hu, Y., … & Cheng, Z. (2020). Clinical features of patients infected with 2019 novel coronavirus in Wuhan, China. The Lancet, 395(10223), 497–506.

12. Khan, M. D., Thi Vu, H. H., Lai, Q. T., & Ahn, J. W. (2019). Aggravation of Human Diseases and Climate Change Nexus. International journal of environmental research and public health, 16(15), 2799. https://doi.org/10.3390/ijerph16152799

13. Kistler, A., Avila, P. C., Rouskin, S., Wang, D., Ward, T., Yagi, S., … & Boushey, H. A. (2007). Pan-viral screening of respiratory tract infections in adults with and without asthma reveals unexpected human coronavirus and human rhinovirus diversity. Journal of Infectious Diseases, 196(6), 817–825.

14. Ksiazek, T. G., Erdman, D., Goldsmith, C. S., Zaki, S. R., Peret, T., Emery, S., … & Rollin, P. E. (2003). A novel coronavirus associated with severe acute respiratory syndrome. New England journal of medicine, 348(20), 1953–1966.

15. Ksiazek, T. G., Erdman, D., Goldsmith, C. S., Zaki, S. R., Peret, T., Emery, S., … & Rollin, P. E. (2003). A novel coronavirus associated with severe acute respiratory syndrome. New England journal of medicine, 348(20), 1953–1966.

16. Kucharski AJ, Russell TW, Diamond C, et al. Early dynamics of transmission and control of COVID-19: a mathematical modelling study. Lancet Infect Dis [Internet] 2020;3099(20):2020.01.31.20019901. Available from: http://medrxiv.org/content/early/2020/02/18/2020.01.31.20019901.abstract

17. Li, Q., Guan, X., Wu, P., Wang, X., Zhou, L., Tong, Y., … & Xing, X. (2020). Early transmission dynamics in Wuhan, China, of novel coronavirus–infected pneumonia. New England Journal of Medicine.

18. Lipsitch, M., Swerdlow, D. L., & Finelli, L. (2020). Defining the epidemiology of Covid-19—studies needed. New England Journal of Medicine.

19. Malik, Y. S., Sircar, S., Bhat, S., Vinodhkumar, O. R., Tiwari, R., Sah, R., … & Dhama, K. (2020). Emerging Coronavirus Disease (COVID-19), a pandemic public health emergency with animal linkages: Current status update.

20. Mandal, S., Bhatnagar, T., Arinaminpathy, N., Agarwal, A., Chowdhury, A., Murhekar, M., … & Sarkar, S. (2020). Prudent public health intervention strategies to control the coronavirus disease 2019 transmission in India: A mathematical model-based approach. The Indian journal of medical research.

21. Maier, B. F., & Brockmann, D. (2020). Effective containment explains sub-exponential growth in confirmed cases of recent COVID-19 outbreak in Mainland China. arXiv preprint 2002.07572.

22. Ming, W. K., Huang, J., & Zhang, C. J. (2020). Breaking down of healthcare system: Mathematical modelling for controlling the novel coronavirus (2019-nCoV) outbreak in Wuhan, China. bioRxiv.

23. Murdoch, D. R., & French, N. P. (2020). COVID-19: another infectious disease emerging at the animal-human interface. NZ Med J, 133(1510), 12–15.

24. Nesteruk, I. (2020). Statistics based predictions of coronavirus 2019-nCoV spreading in mainland China. MedRxiv.

25. Novel, C. P. E. R. E. (2020). The epidemiological characteristics of an outbreak of 2019 novel coronavirus diseases (COVID-19) in China. Zhonghua liu xing bing xue za zhi=Zhonghua liuxingbingxue zazhi, 41(2), 145.

26. Peeri, N. C., Shrestha, N., Rahman, M. S., Zaki, R., Tan, Z., Bibi, S., … & Haque, U. (2020). The SARS, MERS and novel coronavirus (COVID-19) epidemics, the newest and biggest global health threats: what lessons have we learned?. International journal of epidemiology.

27. Perlman S, Netland J. Coronaviruses post-SARS: update on replication and pathogenesis. Nat. Rev. Microbiol. 2009 Jun;7(6):439–50. [PMC free article] [PubMed]

28. Rothan, H. A., & Byrareddy, S. N. (2020). The epidemiology and pathogenesis of coronavirus disease (COVID-19) outbreak. Journal of Autoimmunity, 102433.

29. Voordouw, A. C. G., Sturkenboom, M. C. J. M., Dieleman, J. P., Stijnen, T., Smith, D. J., van der Lei, J., & Stricker, B. H. C. (2004). Annual revaccination against influenza and mortality risk in community-dwelling elderly persons. Jam

30. World Health Organization. Report of the WHO-China Joint Mission on Coronavirus Disease 2019 (COVID-19). 2020.

31. Wrammert J, Smith K, Miller J, Langley WA, Kokko K, Larsen C, Zheng NY, Mays I, Garman L, Helms C, James J, Air GM, Capra JD, Ahmed R, Wilson PC: Rapid cloning of high-affinity human monoclonal antibodies against influenza virus. Nature 2008.

32. Wu, Z., & McGoogan, J. M. (2020). Characteristics of and important lessons from the coronavirus disease 2019 (COVID-19) outbreak in China: summary of a report of 72 314 cases from the Chinese Center for Disease Control and Prevention. Jama

33. Xu, Z., Shi, L., Wang, Y., Zhang, J., Huang, L., Zhang, C., … & Tai, Y. (2020). Pathological findings of COVID-19 associated with acute respiratory distress syndrome. The Lancet respiratory medicine.

34. Yang Yang, Lu Qingbin, Liu Mingjin, et.al. (2020). Epidemiological and clinical features of the 2019 novel coronavirus outbreak in China. MedRxiv, doi: https://doi.org/10.1101/2020.02.10.20021675.

35. Zhang Rongqiang, Liu Hui, Li Fengying, et al. (2020). Transmission and epidemiological characteristics of severe acute respiratory syndrome coronavirus 2 (SARS-CoV-2) infected pneumonia (COVID-19): Preliminarily evidence obtained in comparison with 2003-SARS. MedRxiv, doi: https://doi.org/10.1101/2020.01.30.20019836.

36. Zhao S, Gao D, Zhuang Z, Chong M, Cai Y, Ran J, et al. Estimating the serial interval of the novel coronavirus disease (COVID-19): a statistical analysis using the public data in Hong Kong from January 16 to February 15, 2020. medRxiv 2020c;, doi: http://dx.doi.org/10.1101/2020.02.21.20026559.

